# Effects of the 2019 Mexican health system reform on the Decision-Making Space of federal entities for breast cancer care

**DOI:** 10.1101/2025.05.25.25328320

**Authors:** Beatriz Martínez Zavala, Miguel A. González Block, Mario Salvador Sánchez Domínguez

## Abstract

**Background:** Since 2019, the Mexican health system has undergone a reform aimed at centralizing service provision for the population without social security, replacing the Social Protection System in Health with the decentralized public body IMSS-Bienestar. This operates under two schemes: “adhered” entities, with federal management, and “non-adhered” entities, under state administration. The reform has reshaped intergovernmental relations, affecting local decision-making authority, particularly in the care of chronic diseases such as breast cancer.

**Objective:** To analyze the effects of the 2019 Mexican health system reform on decision-making spaces for breast cancer care across states according to their model of adherence.

**Methodology:** A comparative analysis of decision-making spaces before and after the reform was conducted using Bossert’s Decision Spaces framework. This involved document review and interviews with key stakeholders in one adhered state (Mexico City) and one non-adhered state (Jalisco), using breast cancer as a case.

**Conclusions:** A trend toward centralization was identified, with varying effects depending on the adherence model. In Mexico City, as an adhered entity, centralization encompassed multiple functions, whereas in Jalisco, a non-adhered entity, changes were limited to issues related to service fees and benefit packages. In Mexico City, discrepancies were identified between the de jure and de facto decision-making spaces. Further implementation studies are recommended to explore these discrepancies between formally delegated authority and authority effectively exercised, as well as research linking these spaces to performance and impact indicators, according to the adherence model.

## Introduction

The Mexican health system (SSM) organizes public health services for the population excluded from social security, as well as public health programs, across the three levels of government—federal, state, and municipal—in accordance with the Constitutional framework and the General Health Law (LGS) [1,2]. The Federal Ministry of Health (Secretaría de Salud, SSa) is responsible for governance and the majority of health financing, while health services are delivered by public agencies at either the federal or state level. [3,4] In 2004, the System for Social Protection in Health (SSPH) was implemented to reduce disparities in per capita spending among beneficiaries, lower catastrophic and impoverishing expenditures, incentivize state-level contributions, and improve inter-institutional coordination toward universal coverage [5].

The SSPH operated through the *Seguro Popular* (Popular Health Insurance, SP), a voluntary enrollment scheme regulated and financed by the National Commission for Social Protection in Health (*Comisión Nacional de Protección Social en Salud*, CNPSS) at the federal level and managed by State Regimes for Social Protection in Health (*Regímenes Estatales de Protección Social en Salud*, REPSS) at the state level [5]. The SPSS allowed the participation of accredited private providers to deliver third-level interventions, which were directly financed by the CNPSS through the Fund for Protection against Catastrophic Expenditures (*Fondo de Protección contra Gastos Catastróficos,* FPGC). For first- and second-level services, the State Regimes for Social Protection in Health (REPSS) could engage private providers to address gaps in service availability [6].

In 2020, the SSPH was replaced by the Health for Well-being System (*Sistema de Salud para el Bienestar*, SSB), aimed at reducing out-of-pocket spending, ensuring universal service coverage at all levels, and addressing the corruption issues that affected the SSPH in some states. The reform sought to centralize the provision of health services for the uninsured population under the Institute of Health for Well-being (*Instituto de Salud para el Bienestar*, INSABI), calling on states to transfer their human, material, and financial resources to this institution [7]. INSABI also discontinued agreements with private providers and eliminated enrollment fees or cost-recovery charges for services [8,9]. In 2023, INSABI was replaced by the decentralized public agency Mexican Social Security Institute for Well-being Health Services (*Servicios de Salud del Instituto Mexicano del Seguro Social para el Bienestar*, IMSS-Bienestar), to address the limitations of the previous INSABI [10]. In accordance with the constitutional framework, states had the option to transfer control over their resources to IMSS-Bienestar (‘adherent’ scheme) or to retain it and receive a subsidy from the agency to continue managing service provision (‘not adherent’ scheme) [10,11].

Health system performance is influenced by the relationship between national or federal governments and local or state governments, and by the capacity of this relationship to formulate policies and implement interventions responsive to diverse contexts and populations [12,13]. Comparative analysis of central-local relations requires an assessment of the distribution of power between levels of government and its effect on system performance [13]. Bossert proposed the “Decision Space” (DS) framework to identify the degree of *de jure* or *de facto* autonomy transferred from the national or federal level to subnational levels along five dimensions: financing, service organization, human resources, access rules, and governance. Bossert classified the autonomy of subnational entities as narrow, moderate, or wide, and analyzed the implications for health system equity and efficiency [13,14].

Orozco et al.[15] and González Block et al.[16] applied the DS framework to analyze central-local relations under the SSPH, finding that state governments gained or lost power across various health system functions, with diverse effects on equity and efficiency. Martínez Zavala et al. [17] analyzed the *de jure* changes in decision space during the transition from the SSPH to the SSB, identifying different degrees of centralization across health system functions. The authors emphasized the importance of analyzing the *de facto* centralization following SSB implementation, with a focus on differences in state adherence models [17].

Breast cancer (BC) care has been particularly affected by the changes resulting from MHS reform, especially the centralization of provision and cancellation of contracts with private providers [8,9]. This disease, with high social and economic impact, shows increasing incidence and mortality, associated with limited access to timely detection and treatment services [18]. In the public system, BC care is guaranteed free of charge [2,10]. The institutional response is structured through the Specific Action Program (*Programa de Acción Específico*, PAE) for cancer control, which includes five strategic pillars from primary care to specialized treatment [19], and through the Official Mexican Standard NOM-041-SSA2-2011, which sets the guidelines for prevention, diagnosis, treatment, and epidemiological surveillance [20].

This article analyzes the effects of SSB implementation on the decision space in states that have adhered and not adhered to IMSS-Bienestar. Breast cancer is used as a tracer condition due to its coverage across all levels of care and its sensitivity to the changes introduced by the reform. The comparative analysis focuses on Mexico City and Jalisco, representative cases of the “adherent” and “non-adherent” models, respectively [21,22].

## Methods

The study compares Decision Space (DS) before and after the Health for Well-being System (*Sistema de Salud para el Bienestar*, SSB) between Mexico City, as an “adherent” entity, and Jalisco, as a “non-adherent” entity, based on qualitative analysis of published literature, documentary sources, and key informant interviews. A targeted search of legislative and programmatic health sector documents from the periods 2003–2018 and 2019–2024 was conducted using Google, Google Scholar, and official Mexican government websites. The review included literature on financing, service organization, human resources, access rules, and governance—at both the federal level and the level of the selected entities—across both time periods. In functions or dimensions where breast cancer–specific literature was not identified, broader literature on the public health subsystem for the uninsured population was included.

Mexico City and Jalisco were selected due to their differing political alignments but similar demographic, socioeconomic, and health system characteristics, including installed capacity for breast cancer care. Demographic comparability was ensured by considering variables such as total population (9,209,944 in Mexico City and 8,348,151 in Jalisco), female population (4,805,017 in Mexico City and 4,249,696 in Jalisco), breast cancer mortality rates (19.4 in Mexico City and 22.4 in Jalisco), number of people previously enrolled in the now-defunct SPSS (3.7 million in Mexico City and 4.3 million in Jalisco), and Human Development Index (0.830 for Mexico City and 0.860 for Jalisco) [21,22].

Informants were selected intentionally based on defined criteria. Participants included officials from the federal and state Ministries of Health (*Secretaría Estatal de Salud,* SEDESA), State Health Services (SHS), the IMSS-Bienestar decentralized public agency, and the National Cancer Institute (INCAN), all of whom had knowledge of intergovernmental relations and institutional changes resulting from the reform. A semi-structured interview guide was used, based on the functions of Decision Space and questions regarding the potential effects of the reforms on subnational health system performance (SSM) for breast cancer care. Interviews lasted approximately 60 minutes and were conducted via Zoom between 24/05/2024 and 02/04/2025. All interviews were recorded with participant consent and transcribed verbatim.

Documents and interview transcripts were coded using categories from the Decision Space framework, classifying decision space as narrow, moderate, or wide (Table 1). Decision spaces before and after the reform were mapped for each entity using a matrix with functions and dimensions as rows and decision space levels as columns. This matrix was used to analyze the content of documents and interviews inductively. Quotations were selected through a thorough review of the data matrix, aiming to identify those that best represented emerging patterns in the analysis. Finally, changes in decision space were compared within each entity (pre- and post-SSB) and between the two entities.

**Table 1.**
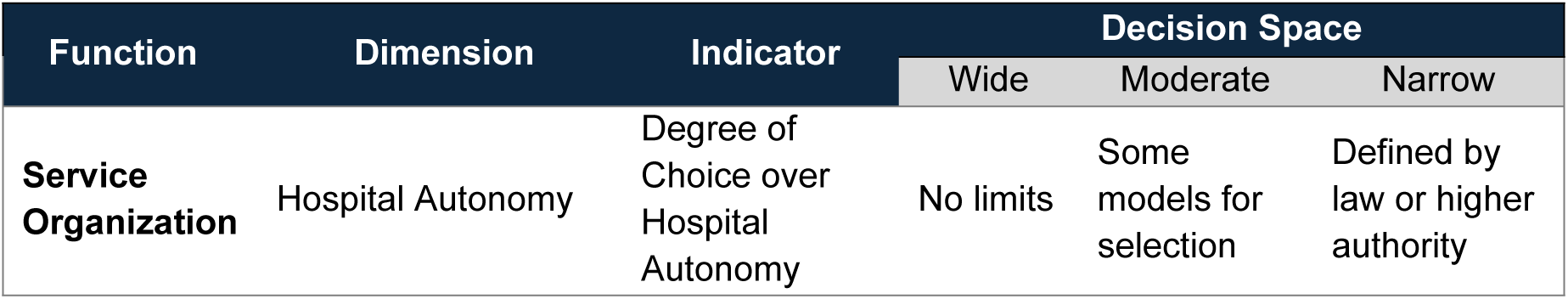
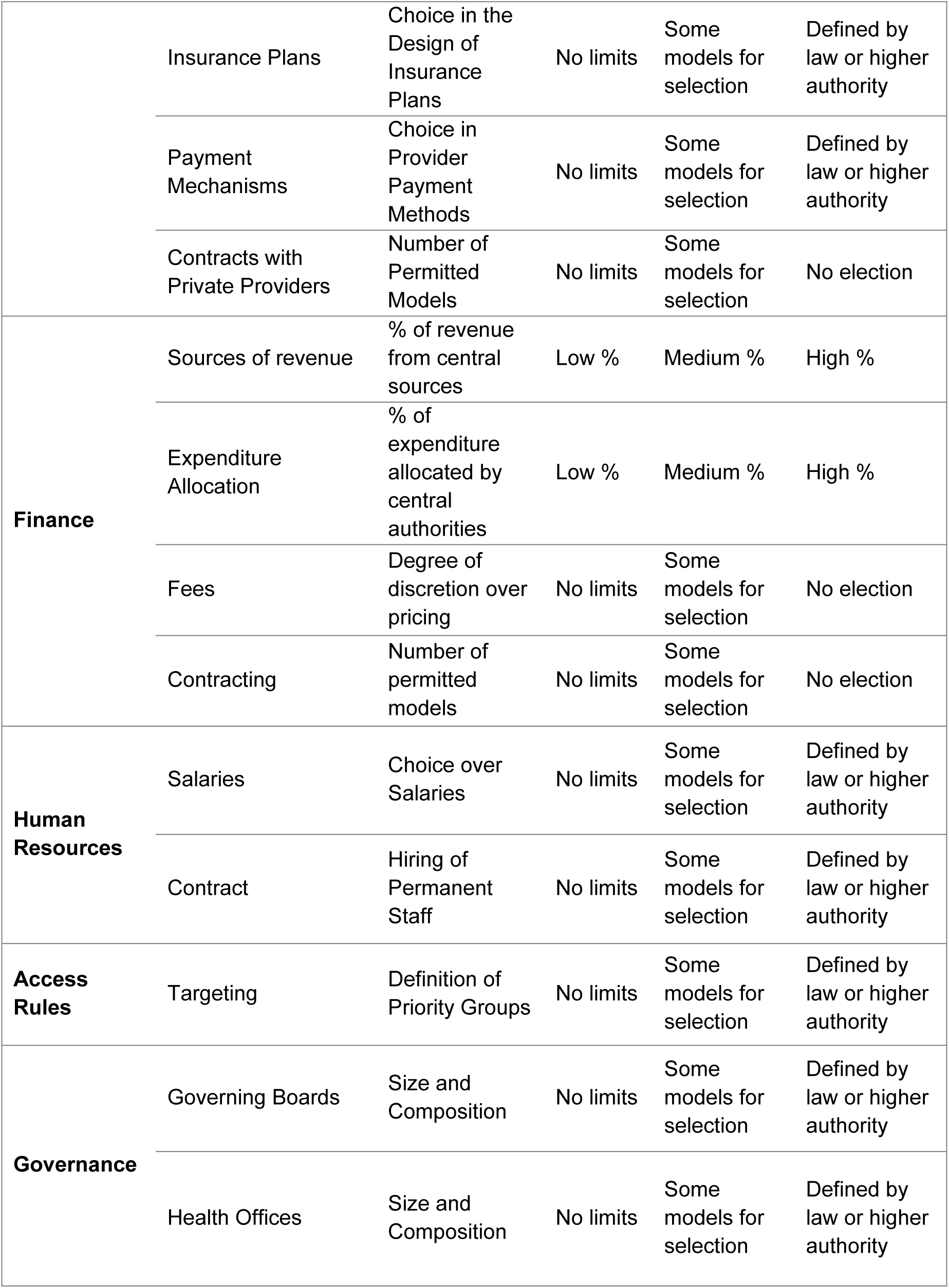
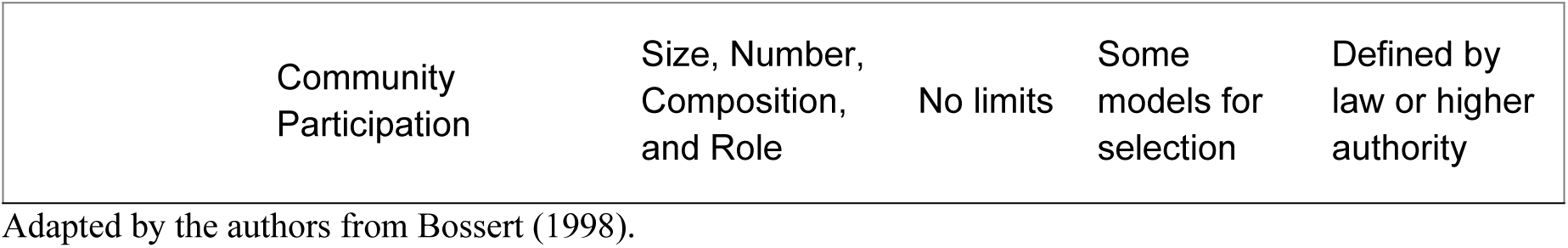
Indicators for Mapping Decision Spaces.

Participation from informants was voluntary, with the study’s objectives explained and written informed consent obtained. Risks were minimized through the protection of personal data and confidentiality in the reporting of results. The study protocol was approved by the Ethics Committee of the National Institute of Public Health (Approval No. CI 1427).

## Results

The profiles of the interviewees are detailed in Table 2.

**Table 2.**
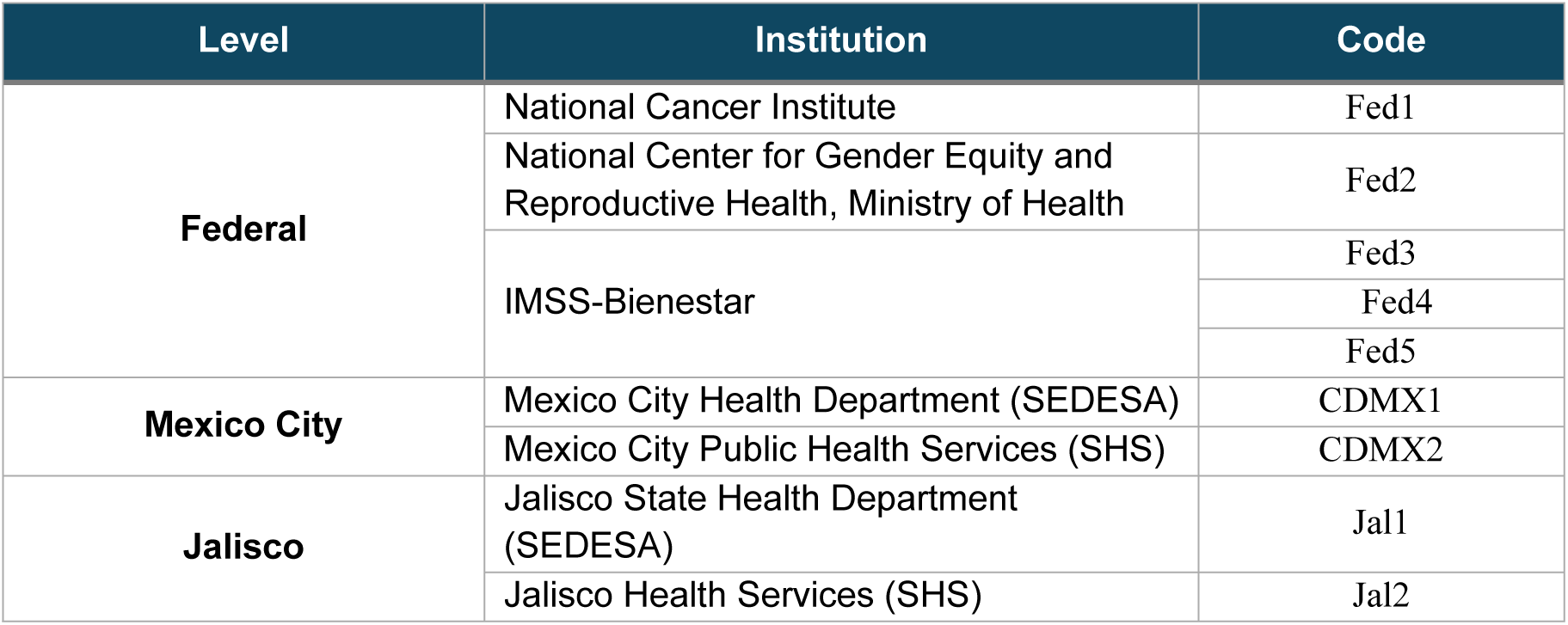
Institution, Profile, and Code of Key Informants Interviewed.

| Level | Institution | Code |
| --- | --- | --- |
| **Federal** | National Cancer Institute | Fed1 |
|  | National Center for Gender Equity and Reproductive Health, Ministry of Health | Fed2 |
|  | IMSS-Bienestar | Fed3 |
|  |  | Fed4 |
|  |  | Fed5 |
| **Mexico City** | Mexico City Health Department (SEDESA) | CDMX1 |
|  | Mexico City Public Health Services (SHS) | CDMX2 |
| **Jalisco** | Jalisco State Health Department (SEDESA) | Jal1 |
|  | Jalisco Health Services (SHS) | Jal2 |

### Service Organization

By 2024, 23 states, including Mexico City (CDMX), had signed agreements with IMSS-Bienestar, while 9 states—including Jalisco—opted to maintain their autonomy [23]. The centralization of services among the “adherent” entities was heterogeneous, as by 2024 the full transfer of assets from some states to IMSS-Bienestar had not yet been completed (Fed4). With the dissolution of the State Health Social Protection Regimes (REPSS), state health ministries integrated funding into their own structures, eliminating the separation between stewardship and financing functions at that level of government.

Insurance plans—defined as the set of interventions covered free of charge—continued to be determined at the federal level. Although the SP and the Universal Catalog of Health Services (*Catálogo Único de Servicios de Salud*, CAUSES) were eliminated, formal coverage was expanded to align with that of social security institutions. The General Health Council (GHC) retained its authority to issue recommendations for the coverage of high-cost diseases [24][25].

In-hospital services for the detection and diagnosis of BC, previously covered by the CAUSES, are now covered by IMSS-Bienestar [2]. Screening and out-of-hospital detection remain under the Women’s Cancer Prevention and Control Program (*Programa de Prevención y Control del Cáncer en la Mujer*, PCCM) [19,26], while treatment—previously funded by the catalog of the FPCG—is now covered by IMSS-Bienestar through the Health Fund for Well-being (*Fondo de salud para el Bienestar,* FONSABI), under the same catalog [26,27].

The agreements with private providers that were previously established by the REPSS were prohibited—a change perceived by stakeholders as a strategy to prevent the misuse of resources and conflicts of interest (Fed2). However, there are expressed concerns regarding the sustainability of this strategy:

> “Let us remember that ISSSTE also tried to provide all services in-house, and it was not cost-effective.” (Fed5).

However, it has not been possible to cancel all outsourced service contracts due to the limited infrastructure available in the states. As a result, a parallel strategy has been implemented to strengthen breast cancer care, based on a situational diagnosis (Fed2).

Private contracts were centralized under IMSS-Bienestar and limited to a portfolio of 21 basic services aimed at ensuring continuity and quality of care. However, in states with greater needs, services outside the catalog may be included in an effort to standardize coverage across the “adherent” entities (Fed5). The state-level IMSS-Bienestar coordinations identify the need for comprehensive services to be contracted and submit these for approval to the Division of Comprehensive and Outsourced Medical Services of IMSS-Bienestar, with the aim of promoting service standardization among states (Fed5).

Through subcontracted services coordinated by IMSS-Bienestar, an integrated care network for BC has been established, encompassing all stages of care, including clinical laboratory, pathology, and imaging services. In the therapeutic domain, mixing centers have been created for the timely and efficient supply of injectable oncology drugs (Fed5).

Stakeholders perceive that with the SSB, coverage was expanded to include cancers that were not covered under the SPSS:

> “Under Seguro Popular, only certain conditions were covered. Today, if not 100%, I would venture to say that at least 85% of oncological conditions are covered.” (Fed1).

In CDMX, healthcare service operations were transferred to IMSS-Bienestar under the management of a State Coordinator—nominated by the Head of Government but employed by IMSS-Bienestar. This entailed the loss of local control over autonomy, contracting, and insurance planning [28]:

> “We had full control because we managed health centers, hospitals, and all early detection and screening activities. Now this changes, because IMSS-Bienestar is taking over control of intramural identification and early detection.” (CDMX1).

Contracts with private providers for breast cancer care were canceled, [8,9,24] although screening services (such as Medibuses) were maintained due to limited installed capacity (CDMX1, CDMX2). SEDESA’s intention is to eliminate these contracts through a program aimed at upgrading and equipping public providers:

> “A shared assessment was carried out across all health centers and INSABI hospitals, IMSS-Bienestar Program, IMSS, and SEDESA. This has already been registered with the Ministry of Finance and approved by FONSABI, which would mean strengthening and renewing equipment.” (CDMX1).

Although contracts and payment mechanisms with private providers are expected to be managed by IMSS-Bienestar, in Mexico City this process has not yet been fully implemented. This is because the IMSS-Bienestar team delegated to the city has not yet consolidated its capacities and structure, and the contracts continue to be designed and signed by SEDESA (CDMX1). Stakeholders note that institutional fragmentation has decreased due to the management efforts of IMSS-Bienestar—specifically, collaboration between SEDESA CDMX and social security institutions aimed at addressing the lack of equipment and infrastructure for conducting biopsies and immunohistochemistry (CDMX1, CDMX2).

The cancellation of private contracts is perceived by CDMX authorities as a recovery of control and resources previously directed to the private sector:

> “Dispensing and medications were subcontracted, and the hospital pharmacy was essentially a private operation over which we had little control. When we took back control and eliminated private pharmacies, we saved 250 million pesos in the first year alone by managing our own dispensing.” (CDMX1).

In Jalisco, with the elimination of the REPSS, a formal institutional restructuring took place, separating the functions of SEDESA and the SHS. Previously, stewardship and service delivery were combined under a single administrative structure. The reform established a differentiated model: SEDESA assumed exclusive normative and oversight functions, while the provision of health services became the responsibility of the decentralized public body (DPA) *Servicios de Salud Jalisco* (SHS) (Jal1, Jal2).

Local authorities noted that, although the federal reform restricted their autonomy by limiting insurance plan design, retaining control over financial and human resources allowed for greater decision-making capacity.

> “There is definitely less room for decision-making, but choosing not to transfer our resources and personnel definitely allows for greater decision-making power across all areas.” (Jal1).

The same perception applies to innovation in treatments:

> “We do have regulations that apply to us, but through public policy proposals, innovations can still be introduced.” (Jal2).

Jalisco’s SEDESA created *Seguro Salud Jalisco*, committing to offer all services included in the IMSS-Bienestar portfolio [29]. The catalog is available online, by level of care, and residents must register online to receive a membership card [29]. Former REPSS functions for promoting health coverage were assumed by SEDESA, while service provision remains under the DPA SHS, Civil Hospitals of Guadalajara, and the Jalisco Cancer Institute [29].

Contracts with private providers were not canceled, and each DPA retains autonomy to select contracting models and payment mechanisms [30]:

> “Each DPA manages its own breast cancer funding. The Civil Hospitals […] perform surgery and have funds for medications; SHS manages policies for the uninsured population and out-of-hospital care; the Cancer Institute is dedicated solely to oncology […] Each institution, within its mandate, manages bidding, contracting, and payments to providers.” (Jal2).

### Finance

The SSB replaced the SSPH’s tripartite financing scheme with a model based on annual budgeting and historical allocations, eliminating affiliation and recovery fees [2,10]. IMSS-Bienestar is funded through resources from Budgetary Program 47 of the Federal Expenditure Budget [31], the federal Fund for Health Services (*Fondo de Aportaciones para los Servicios de Salud,* FASSA), and state-level contributions. ‘Adherent’ states transfer both their state contribution and FASSA funds to IMSS-Bienestar; ‘non-adherent’ states receive direct funding and may agree for IMSS-Bienestar to retain resources for consolidated purchases or infrastructure investment [10].

Resource allocation was also modified. Previously, the budget was allocated to the REPSS based on the number of affiliated individuals, and these funds were invested within each state to provide the covered services. The current centralized process now includes an assessment of installed capacity to allocate resources according to supply-side needs and to reduce disparities between states (Fed2, Fed5).

> “Regardless of the state’s payment capacity […] every state has the possibility to request the same amount of resources; it is only regulated based on what is actually going to be used.” (Fed5).

BC care is financed through IMSS-Bienestar resources for intramural services; the Specific Agreement for the Transfer of Inputs and Allocation of Federal Budgetary Resources (SaNAS)—which replaced the AFASPE—for extramural activities under the PCCM; [32] and the FONSABI through the High-Cost Disease Care Subaccount (*Subcuenta de Atención de Enfermedades de Alto Costo,* SAEAC), which provides annual financing to INCAN and to SEDESAs via in-kind deliveries of medicines and inputs. To access FONSABI resources, SEDESAs must project their annual needs [25,33]. Hospitals must then report the cases treated with these resources to IMSS-Bienestar [25,34],[35]. Additionally, FONSABI includes the Subaccount to Supplement Resources for the Supply and Distribution of Medicines and Other Inputs (*Subcuenta para Complementar los Recursos Destinados al Abasto y Distribución de Medicamentos y Demás Insumos*, SADMI), which allows for the inclusion of high-cost medicines and interventions not listed in the official catalog [25].

In the case of BC, the National Center for Gender Equity and Reproductive Health (*Centro Nacional de Equidad de Género y Salud Reproductiva*, CNEGSR) continues centralized procurement of strategic inputs for PCCM implementation but disburses funds to states when consolidated purchasing is not undertaken—regardless of adherence to the SSB. Stakeholders view centralized funding for BC as a means of enhancing evaluation in both allocation and implementation:

> “The benefit of being able to centralize again is precisely having much greater control and evaluation.” (Fed2).

However, the shift toward in-kind resource transfers is perceived as a limitation for innovation in infrastructure and equipment:

“When the hospital received financial resources directly, it enabled the development of infrastructure […] because, in the end, all the funds were in a single pool.” (Fed1).

In Mexico City, prior to the reform, 82% of the total SP funding came from federal sources and 18% from state sources (Table 3). The FASSA funds were received with specific earmarks for disbursement in defined categories. Additionally, 94% of the resources for women’s cancer care were allocated in kind by the federal government [36][37]. After the reform, there was an increase in the state contribution, which now represents 59% of the total resources of IMSS-Bienestar. However, the allocation of these resources became centralized with the transfer of both the state contribution and FASSA funds to IMSS-Bienestar for their management [28]. Additionally, the federal government retained control over PCCM resources, allocating 96% in-kind [38].

**Table 3.**
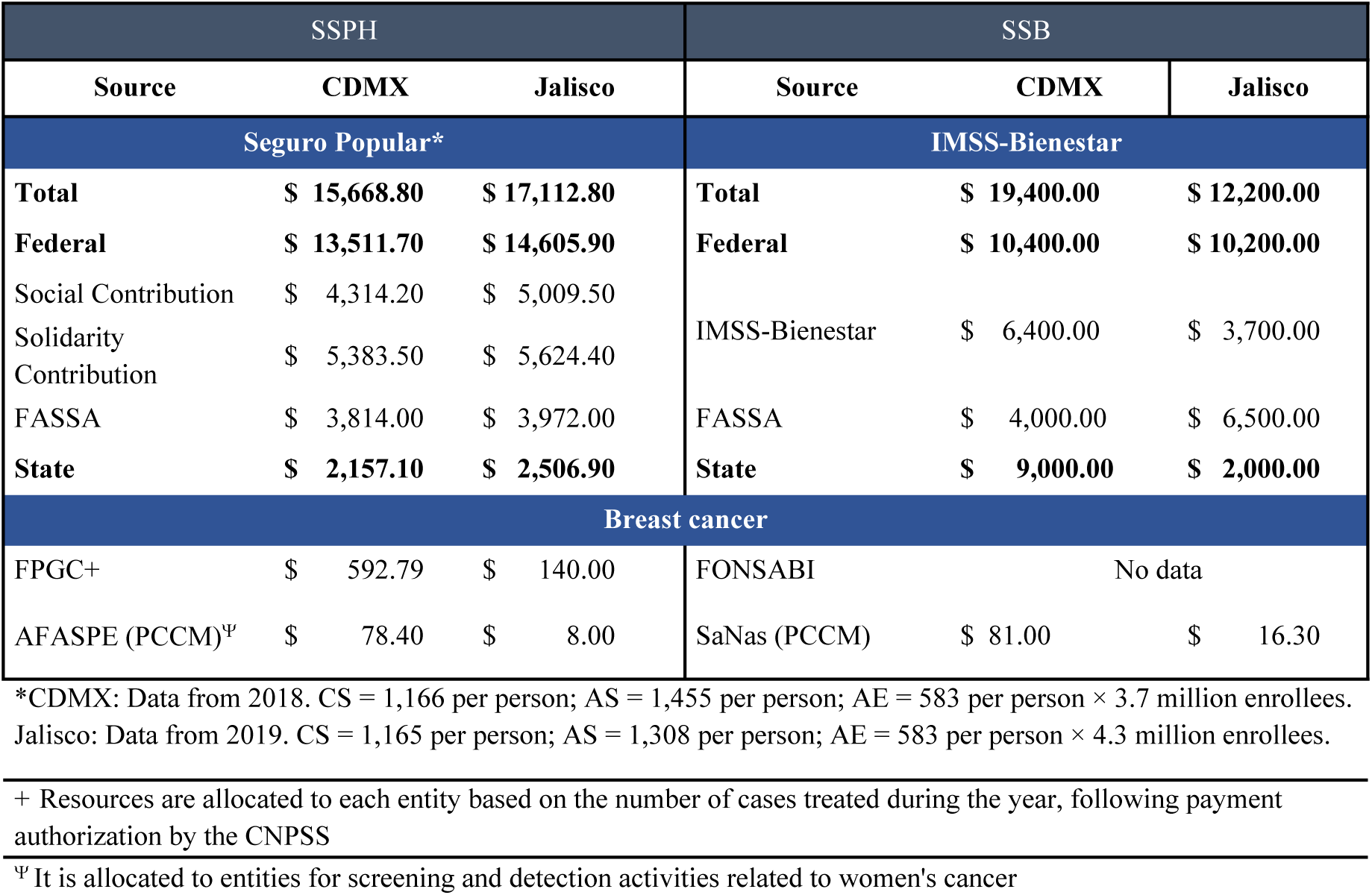
Financing in CDMX and Jalisco Before and After the Reform.

In CDMX, the centralization of resources and procurement under IMSS-Bienestar is perceived as a barrier to timely responses for patients with specific needs:

> “[Before] we were able to respond quickly to what we call the patient profile. [Now] it’s more difficult because everything has to be requested through IMSS-Bienestar […] and also with medicines, because the directive was to centralize everything through Birmex.” (SEDESA, CDMX).

In Jalisco, before the reform, 81% of SP funding came from federal sources (Table 3). All FASSA resources were earmarked for specific actions, and 99% of PCCM funds were received as direct financial transfers [39]. After the reform, Jalisco received 65% of IMSS-Bienestar resources, and FASSA transfers [40], while 98% of PCCM resources were received in-kind [40]. Although, after the reform, federal contributions for individual care decreased from $10,634 million to $3,700 million in this entity, the federal contributions from FASSA and PCCM increased by 1.6 and 2.03 times, respectively, compared to 2018. This seems to compensate for the differences in federal contributions through IMSS-Bienestar between entities, with the total difference between entities being due to the contribution from the non-adherent entity (Table 3).

Jalisco retained the REPSS’s resource allocation authority, now exercised through SEDESA under federal spending limits for medicines, inputs, and personnel—similar to those imposed under the SSPH: up to 50% of transferred federal funds may be allocated to personnel directly involved in healthcare delivery, with the remainder designated for medical unit operations, as well as the acquisition and distribution of medicines and inputs. Furthermore, 20% of federal resources must be used for disease prevention and health promotion activities [41].

Maintaining this purchasing capacity is viewed positively in the state:

> “We already had the capacity to procure oncology medicines, and that prevented the shortages we saw in other parts of the country.” (Jal2).

The entity acknowledges the administrative benefits of centralizing resources for the PCCM; however, it considers that this complicates the receipt of supplies tailored to its needs:

> “Many times [the suppliers] do not deliver the medication with the agreed specifications or within the timeframes established in a centralized contract.” (Jal2).

DPAs retained the capacity to charge recovery fees for certain complementary services in accordance with regulations and fee schedules:

> “In Jalisco, various DPAs may apply recovery fees. The only one prohibited from doing so is *Servicios de Salud Jalisco* […] but other DPAs—like the cancer institute, civil hospitals, and municipal medical services—do charge. By law, there are conditions under which fees are not allowed […] But for activities unrelated to specific diseases, fees can be charged.” (Jal2).

### Human Resources

Historically, health personnel in SHS have been hired through federal or state positions, negotiated with federal unions. [42] IMSS-Bienestar integrated staff holding federal contracts and hires new personnel under federal positions, assigning them to work in the adhering states [2,10]. IMSS-Bienestar has also redistributed personnel to standardize staffing densities across localities and initiated the process of rehiring with tenured positions (*basificación*) state-level personnel who were previously hired on a fee-for-service basis under the SSPH [43]. However, the existence of multiple employment schemes and salary disparities between levels of government has added complexity to the hiring and regularization processes under IMSS-Bienestar:

> “It has not been simple. Because state legislation for former state employees differs from what the federal government is offering, and that leads to a negotiation process. And that will take time.” (Fed5).

In CDMX, health personnel hiring was centralized, and positions in health services, public health, and governance previously under SEDESA were transferred to IMSS-Bienestar [28,33]. Although the staff is hired and coordinated by IMSS-Bienestar, SEDESA is responsible for selecting them (CDMX1, CDMX2). The process of re-hiring with permanent positions was also implemented:

> “Now, under IMSS-Bienestar, those with precarious contracts were prioritized for hiring. Most of them have now been contracted, as long as they were working in healthcare units.” (CDMX1)

In Jalisco, there were no changes in the hiring process for human resources, and the regularization of fee-for-service staff continued with state resources [34]. However, personnel hired with IMSS-Bienestar funds must be approved by the federal state:

> “As a ‘non-adherent’ state, each DPA handles its own hiring. The state carries out the process… [IMSS-Bienestar] tells you what criteria they must meet and performs a validation of the candidates you propose.” (Jal2).

### Access Rules

The definition of priority groups for coverage remains under the MoH, designating those without social security as beneficiaries of IMSS-Bienestar or SHS, depending on the collaboration model, while prioritizing groups with greater marginalization [2,44]. IMSS-Bienestar eliminated the affiliation process and the insurance policies previously implemented under the SSPH, now requiring only a valid official identification for individuals to receive care at medical facilities. Screening for BC is regulated by the MoH for specific age and risk groups, based on the Official Mexican Standard 041-SSA-2-2011 [20], the PAE [19], and the GHL, with no changes from the SSPH framework [2].

IMSS-Bienestar implemented a beneficiary registration system at healthcare units and an optional online registration platform, which includes the issuance of a beneficiary ID card [2,10]. In adhering states, SEDESAs are now responsible for promoting beneficiary registration [33]. In Mexico City, SEDESA is committed to supporting registration through the IMSS-Bienestar platform [28].

In Jalisco, SEDESA committed to providing free healthcare services to state residents without social security, promoting access to services through the *Seguro de Salud Jalisco*, which follows the coverage framework established by IMSS-Bienestar [45]:

> “We continue to uphold free services, obviously for our target population […] The *Seguro Salud Jalisco* project has been developed, which includes a benefits catalogue, but it is the same as the one used in all states.” (Jal1).

### Governance

The implementation of the SSPH was overseen by the MoH, supported by the National Health Council (*Consejo Nacional de Salud*, CNS), the body responsible for planning, programming, and evaluating actions undertaken by the states and their respective State Health Councils. [46,47],[48] The MoH retained its regulatory role under the new SSB, while the CNS was renamed the National Health Council for Wellbeing (*Consejo Nacional de Salud para el Bienestar*, CONASABI) [2].

CONASABI has played a key role in the governance of strategies such as the replacement of mammography equipment led by CNEGSR, ensuring state-level commitment to equipment maintenance and productivity:

> “It was a way to compel the states and health secretaries—who are the ones signing at the CONASABI—to commit to something they can no longer overlook, regardless of adherence model.” (Fed2).

Under the SSPH, the PCCM was implemented by state SEDESAs in coordination with the CNEGSR. Under the SSB, the National Public Health Service (NPHS) was established to coordinate all health promotion and disease prevention activities under federal leadership [49,50]. This centralization of responsibilities is perceived as a way to strengthen public health:

> “Now the National Public Health Service sits at the table […] They are not involved in direct operations. They will be responsible for supervising that the operational teams do things correctly. And it’s good to have them at the table because they are essentially our counterpart.” (MoH).

IMSS-Bienestar is governed by a board of directors appointed by the president of the republic, who also appoints its general director. The State Coordinations of IMSS-Bienestar are subordinated to the General Directorate without independent financing (Fed4)[51].

In the “adhered” states, IMSS-Bienestar Coordinations are responsible for configuring the care networks for BC care, while SEDESAs retain a regulatory and supervisory role [28]. However, SEDESAs are seen as struggling to adapt to their new functions:

> “What I’ve seen is that the State Health Secretariats still haven’t figured out—or haven’t been given the opportunity—to redesign their structure, purpose, and objectives.” (Fed5).

As part of the transition to the SSB, the existing Sanitary Jurisdictions are being replaced by Districts for Health and Wellbeing (*Distritos de Salud para el Bienestar*, DSB). The coordination among DSBs will be managed by a state-level coordination and regulatory unit headed by a representative of the MoH in each state [52][53]. This implies a reorganization of the populations under responsibility for first-level care and collective health actions, which include screening, diagnosis, and education for the prevention and detection of BC [19].

The new DPA IMSS-Bienestar, created under the SSB, did not replace or integrate the long-standing IMSS-Bienestar Program run by IMSS with federal funds to serve marginalized populations. While the segmentation persists, the closer relationship between the two programs has enabled collaboration to improve services and infrastructure [54].

Community participation in health has remained largely unchanged. According to the GHL, institutions and local governments must promote citizen involvement in decision-making and service improvement and encourage social oversight to ensure transparency. Users retain the right to file complaints, which must be addressed in a timely manner, and participation does not condition access to health services [2,10,33,45].

The Health Care Model for Wellbeing (*Modelo de Atención a la Salud para el Bienestar*, MAS-Bienestar) proposes the gradual formation of various social participation bodies, including local forums, district assemblies, municipal intersectoral committees, municipal health networks, and both virtual and population-based initiatives. [55] In both states adhering to the DPA IMSS-Bienestar and those served by the IMSS-Bienestar Program, the initiative *La Clínica es Nuestra* (The Clinic is ours) has been implemented. This program focuses on strengthening primary care infrastructure through community committees [54,56].

In Mexico City, SEDESA remains the chief regulator of the state health system and supervises IMSS-Bienestar’s performance [28,33]. SEDESA also established the State Inter-Institutional Health Sector Command (*Comando Estatal Interinstitucional del Sector Salud*, CEISS) to coordinate the reform process:

> “While the City Health Council is a deliberative and consultative body, the CEISS has a far more operational task—execution and decision-making to manage the entire system. IMSS, ISSSTE, and Pemex are service providers, and the Secretary of Health chairs the council as the system’s regulator.” (CDMX1).

In Jalisco, the boards of directors of the DPAs and their decision-making authority to configure healthcare networks, manage resources, and provide services remain in place [34]. The 2019 functional separation, which included the establishment of the Directorate of Public Policy within SEDESA to enhance autonomous health policy formulation, is viewed as beneficial for governance:

> “Having this model reduced the impact of the reforms because there is a body responsible for directing and formulating health policy, which also allows us to anticipate risks and prepare responses using state resources.” (Jal1).

Health authorities in Jalisco view CONASABI as an instrument for governance in BC:

> “CONASABI […] determines the budget and what I will receive in cash and in-kind, and sets targets for a specific number of mammograms or screenings.” (Jal2).

However, the lack of clarity in the collaboration agreements to assign responsibilities between the federal and local levels in non-adherent entities became evident in Jalisco as limitations in planning and the receipt of supplies (Jal1, Jal2).

## Discussion

This research analyzed the *de jure* and *de facto* effects of the 2019 reform to the SSM on decision spaces in two entities with different adhesion models, highlighting a trend toward centralization.

After the reform, the autonomy of providers in the ‘adherent’ state was reduced to a narrow decision space. However, through collaboration between state and federal levels, IMSS-Bienestar promoted agreements to expand provider networks through inter-institutional cooperation. While centralization limited the autonomy of institutions for innovation, this did not mean a lack of innovation for the attention of BC within the SSB, as evidenced by the replacement of mammography machines with digital ones, the launch of new guidelines for prevention, screening, diagnosis, treatment, control, and surveillance of BC in Mexico, and the inclusion of new BC therapies at the national level during the SSB [57,58].

Under the SSPH, REPSS had moderate spaces to subsidize public providers and contract private ones. After the reform, the decision space for contracting private services in ‘adherent’ state was narrowed, while the ‘non-adherent’ state remained unchanged. The contracting of private services managed through IMSS-Bienestar could reduce deficiencies in care, particularly in rural areas, through networks based on patient navigation and proximity. However, strengthening state capacity in lagging entities may require a significant investment of time and resources during the initial implementation phase of the reform.

Decisions about insurance plans remained centralized in both states before and after the reform. REPSS did not have the capacity to modify the CAUSES, but they had wide space to offer services outside the catalog with state resources [17]. With the SSB, decision spaces for defining insurance plans were narrowed in both types of entities, as they committed to offering all first- and second-level services following federal guidelines. However, each third-level DPA in Jalisco has the margin to define its portfolio based on its resolutive level. The broad decision-making spaces to modify insurance plans with state resources under the SSPH may have increased inequity between entities, as variations in BC care indicators were documented between regions of the country, showing an inverse relationship between marginalization index and process indicators [59]. Thus, the restriction of decision spaces in the organization can promote greater homogeneity between federal entities, although at the same time, it reduces the state’s capacity to adapt health models to the needs of the population [13].

Table 4 summarizes the changes in decision spaces in the health subsystem for the population without social security.

**Table 4.**
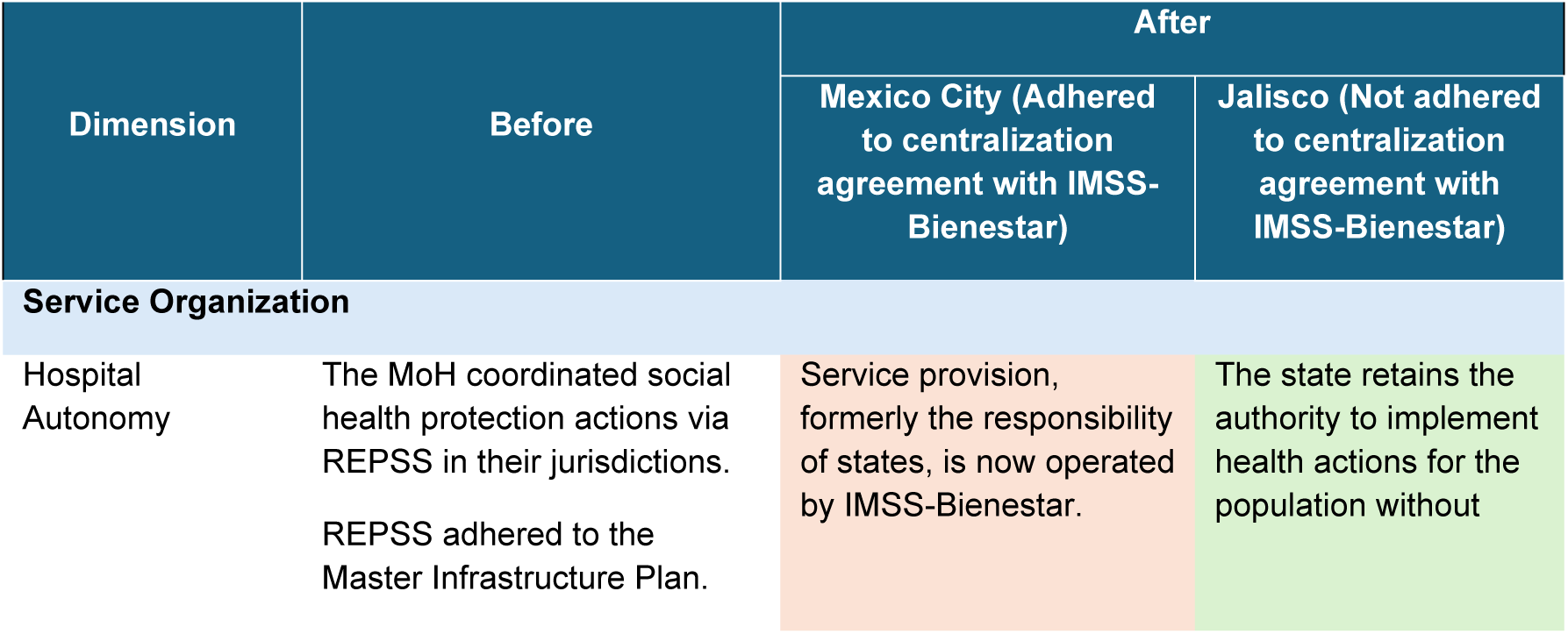
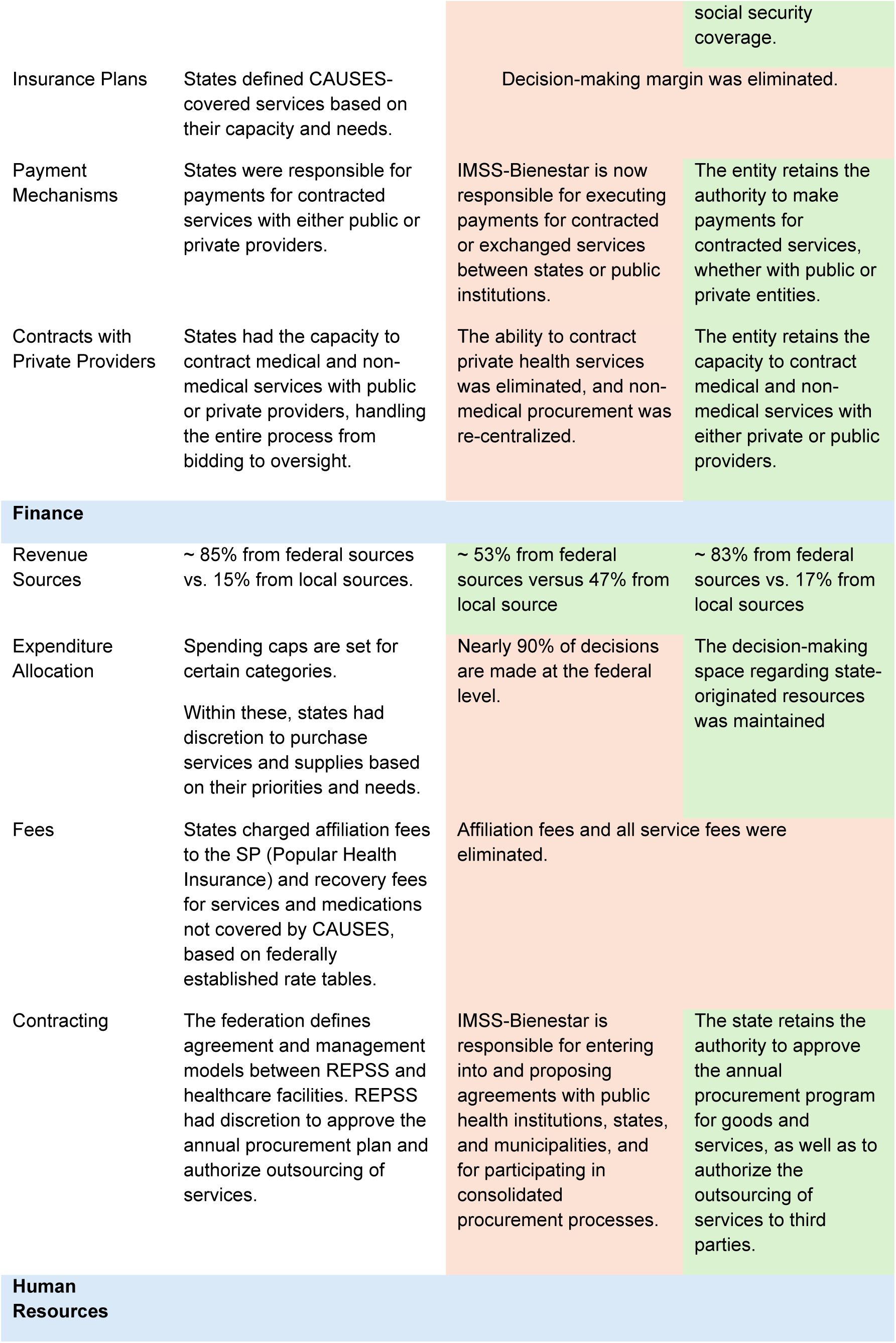
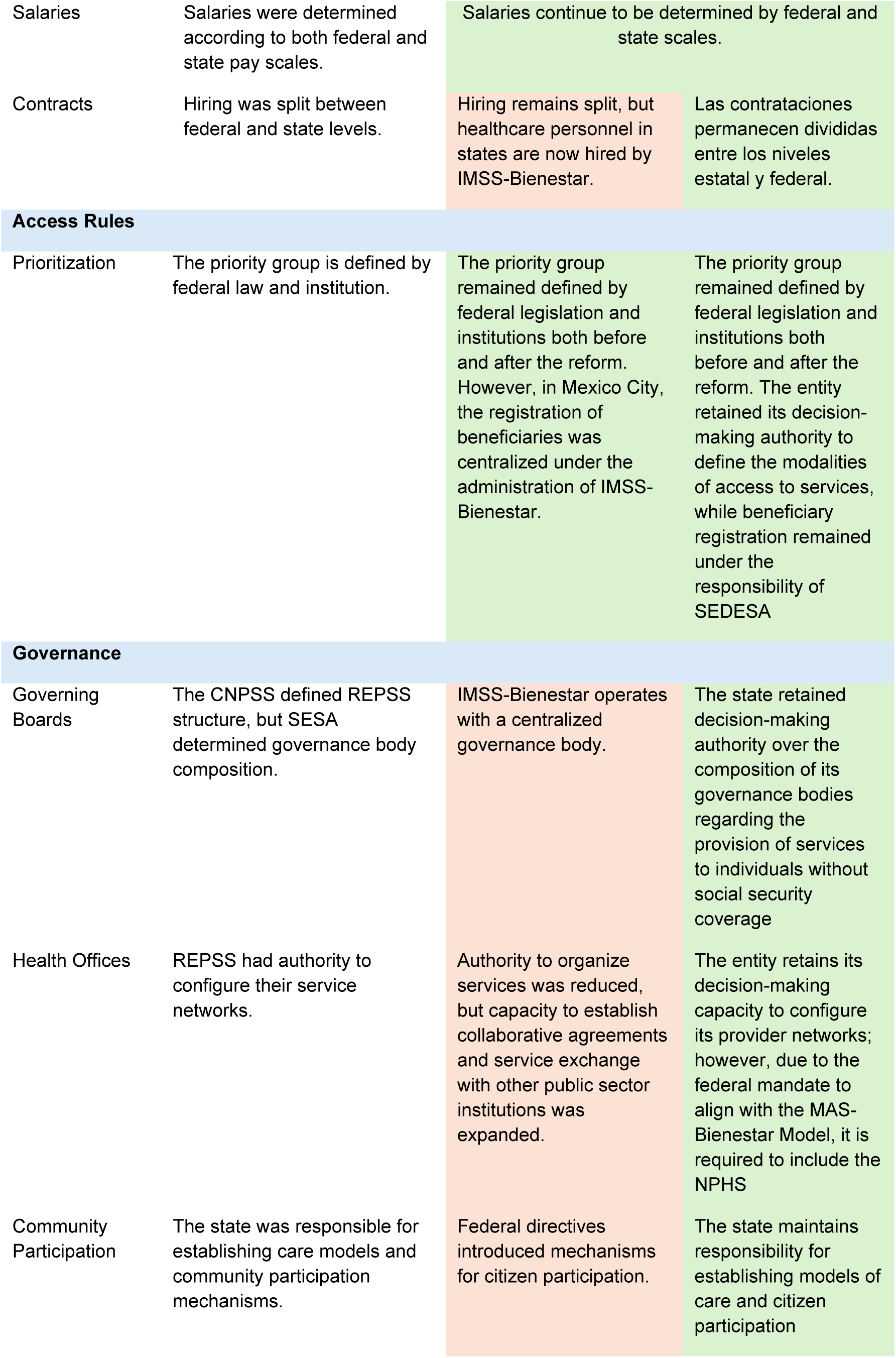
Decision spaces for financing before and after the SSB, CDMX and Jalisco. Centralization is marked in orange, and areas where no changes occurred are marked in green.

Own elaboration.

In CDMX, DS for contracting services with private providers and defining payment mechanisms were narrowed. However, due to the limited experience of IMSS-Bienestar, SEDESA maintained this control and advised IMSS-Bienestar until 2024. In contrast, Jalisco’s spaces for these dimensions remained moderate, allowing institutional reorganization with a commitment to adopting MAS-Bienestar and the NPHS.

Before the reform, the DS for financing first- and second-level BC care were moderate, with federal funding in cash and kind for extramural screening activities from the CNEGSR program and state and federal funding from the SSPH for the execution of intramural activities covered by CAUSES. Although screening and diagnosis of BC were exempt from fees, entities charged affiliation fees to the SP and recovery fees for diagnostic and complementary processes not included in CAUSES. In the adhering state, these spaces were narrowed, with state contributions centralized and receiving personnel, supplies, and medications from IMSS-Bienestar in kind. The non-adhering state maintained moderate DS for financing, receiving federal resources in cash and kind from IMSS-Bienestar for first- and second-level BC care, though resource allocation was centralized through the CNEGSR program. Only in the “fees” dimension did the decision space change to narrow, as affiliation and recovery fees were canceled.

The reform deepened the centralization of financing for third-level BC care in both ‘adherent’ and ‘non-adherent’ entities by replacing direct payment for service packages with a scheme based on annual programming, primarily in kind, through the FONSABI. Although no reports were identified to compare resource allocation before and after the reform, interviews and normative documents pointed to FONSABI as a primary source of funding [35,60].

The centralized purchase and adjudication of medicines by Birmex has received recognition from international organizations [61], however, in practice, the strategy has not resulted in sufficient supply and distribution [62,63]. Furthermore, the centralized purchase by Birmex was perceived in CDMX as a restriction that affects the capacity for medical response, while in Jalisco, delays in medicine deliveries were observed in the first year of the reform. These limitations highlight the importance of strengthening cooperation between entities and IMSS-Bienestar to delineate competencies and optimize mechanisms for resource adjudication and distribution.

Prior to the reform, the entities had a moderate DS for hiring personnel through state positions financed by SSPH and their own resources in administrative and research functions. However, the federation exercised control through the definition of spending caps, salary scales, and approved hiring profiles. After the reform, in adhering entities, this DS was narrowed as state positions and new hires were transferred to IMSS-Bienestar. In non-adhering entities, the situation remained unchanged.

In both entities, the priority group remained defined by a federal law and institution both before and after the reform. However, in Mexico City, the beneficiary registry was transferred to IMSS-Bienestar for administration, while in Jalisco, the registry remains under the responsibility of SEDESA.

Decision spaces in governance in the adhering state shifted from moderate to narrow, especially with the transfer of network coordination to IMSS-Bienestar. In this state, the implementation of state governance bodies for reform implementation in collaboration with the federal level was favored. In the non-adhering state, DS for governance, configuration of health offices, and boards of directors remained unchanged.

The coexistence of two centralization models by the SSB, ‘adhering’ and ‘non-adhering’, generated greater heterogeneity in the relationships between the central government and the states compared to the more homogeneous DS observed under the SSPH. The evidence suggests that local policies can have differentiated effects on BC care and survival [64], highlighting the need for cancer control plans that address local needs within a national framework [65]. Thus, it is crucial to foster coordination between levels of government, promoting joint planning.

In both periods, the PCCM implementation has been highly vertical, with central financing and planning and little decision space for entities. The integration of NPHS into the activities of screening and detection within PCCM exacerbates centralization. The SSB’s proposal to integrate federal programs under NPHS aims to reduce duplicated activities and optimize resource use [66]. Although this body has not been established in the non-adhering state, MAS-Bienestar indicates that its implementation will be nationwide [49]. Interviews suggest that centralization is perceived as a useful mechanism for unifying administrative processes and strengthening inter-institutional collaboration. In this way, NPHS could play a crucial role in integrating federal programs and ensuring the continuity of BC care [66], especially in the face of changes in assurance schemes and labor conditions, which have been shown to disrupt access to care for chronic diseases in Mexico [67].

This study observed that entities increased their financial contribution after the reform. A study by González Block et al. [60], which compared a centralized state with a decentralized one during Mexico’s first decentralization period, found that decentralized entities paradoxically depended more on central financing than centralized ones. These findings suggest that central control predominates in Mexico’s health sector. Even under decentralizing policies such as SSPH, the decision spaces identified were mostly moderate. These results align with previous studies that highlight limited DS for local level and high central control, particularly in the financial realm [15,16]. The findings of this study confirm differences in the effects of the 2019 MHS reform according to the adhesion model as proposed by Martínez et al. [17].

Although the findings are not generalizable to other entities or health conditions, the strategic selection of representative cases of the models and using BC as a tracer allows for observing changes in all three levels of care within the MHS. Given that more than 70% of entities in Mexico have adhered to the new model [23], it can be inferred that DS in most of the country may be similar to those in the adhering state analyzed. The study addresses BC as a public health issue that reflects historical barriers to access for the population without social security, seeking to promote quality and equity in services.

This research is the first in Mexico to examine the impact of the 2019 health reform on decision spaces and the recentralization of services using a comparative approach. Its relevance lies in the modifications to the GHL and their effect on inter-institutional dynamics, offering a replicable conceptual framework and a methodology suitable for middle-income countries.

This study identified differences between *de facto* and *de jure* decision spaces. In the ‘adherent’ entity, the contracting of private services for breast cancer screening was not cancelled, nor has the management of contracts with providers been transferred to IMSS-Bienestar. Furthermore, neither entity has fully implemented the SNSP. These discrepancies between *de facto* and *de jure* decision spaces may be attributed to the recent transition of the health services model towards a new framework, as well as to the implementation process itself, suggesting that the observed variations do not necessarily reflect differences in objectives or local barriers. It is recommended to deepen the implementation analysis to identify specific barriers and facilitators.

Mexico City and Jalisco present some demographic contrasts, with the former being the most densely populated entity in the country, with 6,163 inhabitants per km², while Jalisco has 106 inhabitants per km²; thus, rural population in Mexico City represents 0.7% of the total, while in Jalisco it accounts for 12%. These differences affect access to specialized health services, such as those required for BC care. However, they were chosen to represent opposite coordination models, ensuring reasonable structural and demographic comparability.

The key informants interviewed represented federal, state, and institutional levels of interest for the study. Their profiles demonstrated experience in key positions both before and after the reform, and regardless of the institution, level, or entity they represented, the informants expressed both favorable and critical opinions of both periods. Although the methodology applied avoided including opinions not supported by the review or the rest of the interviews, it is suggested to interpret the cited statements critically.

Due to the recent implementation of the reform, its impact on performance cannot yet be assessed. In the medium and long term, it is suggested to link the decision spaces derived from the reform with indicators of timely access to diagnosis, continuity of treatment, and health outcomes related to breast cancer, both before and after the reform, under the two coordination models with IMSS-Bienestar.

## Conclusions

A trend toward centralization was identified in both entities analyzed, although with differentiated effects. In Mexico City, the ‘adherent’ entity, most functions were narrowed, while in Jalisco, the ‘non-adhered’ entity, the changes were limited to narrow spaces for fee collection and the definition of service packages.

Implementation research is recommended to analyze the differences between the identified *de jure* and *de facto* decision spaces, as well as research on decision spaces with performance and impact indicators, considering the modalities of adhesion.

## Data Availability

The information provided by participants in this study was collected under a confidentiality agreement that restricts its use exclusively to this research. Therefore, full transcripts or detailed excerpts cannot be shared publicly without violating this agreement. Relevant quotations that support the analysis are included within the body of the manuscript. Additional data may be made available upon reasonable request, subject to ethical approval and confidentiality considerations.

## Notes

### Competing Interest Statement

The authors have declared no competing interest.

### Funding Statement

The author(s) received no specific funding for this work.

### Author Declarations

Participation of informants was voluntary. The study’s objectives were explained, and informed consent was obtained. Risks were minimized through protection of personal data and confidentiality in the dissemination of results. The research protocol was approved by the Ethics Committee of the National Institute of Public Health (No. CI 1427).

